# Socioeconomic, racial and ethnic differences in patient experience of clinician empathy: Results of a systematic review and meta-analysis

**DOI:** 10.1101/2020.07.08.20148858

**Authors:** Brian W. Roberts, Nitin K. Puri, Christian J. Trzeciak, Anthony J. Mazzarelli, Stephen Trzeciak

## Abstract

**Introduction:** Empathy is essential for high quality health care. Health care disparities may reflect a systemic lack of empathy for disadvantaged people; however, few data exist on disparities in patient experience of empathy during face-to-face health care encounters with individual clinicians. We systematically analyzed the literature to test if socioeconomic status (SES) and race/ethnicity disparities exist in patient-reported experience of clinician empathy.

**Methods:** Using a published protocol, we searched Ovid MEDLINE, PubMed, CINAHL, EMBASE, CENTRAL and PsychINFO for studies using the Consultation and Relational Empathy (CARE) Measure, which to date is the most commonly used and well-validated methodology for measuring clinician empathy from the patient perspective. We included studies containing CARE Measure data stratified by SES and/or race/ethnicity. We contacted authors to request stratified data, when necessary. We performed quantitative meta-analyses using random effects models to test for empathy differences by SES and race/ethnicity.

**Results:** Eighteen studies (n=9,708 patients) were included. We found that, compared to patients whose SES was not low, low SES patients experienced lower empathy from clinicians (mean difference= -0.87 [95% confidence interval -1.72 to -0.02]). Compared to white patients, empathy scores were numerically lower for patients of multiple race/ethnicity groups (Black/African American, Asian, Native American, and all non-whites combined) but none of these differences reached statistical significance.

**Conclusion:** These data suggest an empathy gap may exist for patients with low SES. More research is needed to further test for SES and race/ethnicity disparities in clinician empathy and help promote health care equity.

**Registration (PROSPERO):** CRD42019142809

## Introduction

Empathy is sensing and detecting another’s emotions, resonating with their thoughts and feelings, and sharing and understanding their perspective. In health care, empathy is a vital clinical competency – an emotional bridge that drives compassionate care for patients.^1^ As such, empathy is essential for high quality health care. Numerous studies published in the literature show that clinician empathy is associated with better patient outcomes across many different medical conditions.^2–14^

Health care disparities are meaningful differences in health care quality that exist between population groups (e.g. race, ethnicity, gender, sexual orientation) not explained by variation in patient preferences, health care needs, or treatment guidelines, and often linked with socioeconomic disadvantage.^15^ Although clinicians ought to have empathy for all patients, it is possible that disparities exist in empathy from clinicians. On a systems level, disparities in access to health care may be rooted in a societal lack of empathy for disadvantaged persons (e.g. institutionalized racism). Health care disparities occurring at the point of care with individual patients may be due to clinician bias (e.g. implicit or unconscious bias), and this may involve a lack of empathy. Examples include inadequate analgesia for Black/African American and Hispanic/Latino patients with painful conditions,^16–19^ inappropriately low use of cardiac catheterization for Black/African American patients with possible acute myocardial infarction,^20^ and clinicians’ false assumptions that Black/African American patients will have poor adherence to treatment recommendations,^21^ among many others. Although some studies have reported that Black/African American patients and Hispanic/Latino patients have hospital experiences that are not worse than those of white, non-Hispanic patients,^22,23^ other studies have shown that race/ethnicity and SES differences exist in patient satisfaction with clinicians,^24,25^ possibly due to lower quality interpersonal interactions and clinician-patient relationships.^26,27^ However, few data exist on SES and race/ethnicity disparities in patient experience of clinician empathy (e.g. *interpersonal* racism), specifically.

The Consultation and Relational Empathy (CARE) Measure is to date the most commonly used and well-validated methodology (i.e. proven reliability, internal validity and consistency ^28^) for measuring clinician empathy from the patient perspective.^29,30^ The Electronic Supplementary Material contains the ten questions that comprise the CARE Measure **(Supplementary Material 1)**. On a 40-point scale (range 10 [lowest] to 50 [highest]), the instrument measures a patient’s assessment of the empathy of a clinician, for example listening and understanding, being interested in the patient as a whole person, and showing compassion.

We hypothesized that low SES patients (compared to not low SES patients) and Black/African American and Hispanic/Latino patients (compared to white, non-Hispanic patients) report lower empathy from clinicians. We aimed to test this hypothesis by conducting a systematic review and meta-analysis of all published studies containing data for patient assessment of clinician empathy using the CARE Measure.

## Methods

### Protocol and registration

We developed a protocol for this systematic review and meta-analysis and published it previously.^31^ The protocol was developed in accordance with the Cochrane Handbook,^32^ and reported in accordance with the Preferred Reporting Items for Systematic Reviews and Meta-Analysis Protocols (PRISMA-P) statement.^33^ We report our results in this manuscript in accordance with PRISMA and the Meta-analysis of Observational Studies in Epidemiology (MOOSE) guidelines.^34,35^ The MOOSE checklist is uploaded as a separate file in the supplementary material. We prospectively registered this systematic review in the PROSPERO international prospective register of systematic reviews (CRD42019142809). This systematic review did not collect individual patient-level data and therefore did not require ethical approval.

### Eligibility criteria

We considered any study in which patients rated their clinicians’ empathy using the CARE Measure to be eligible for potential inclusion. Our inclusion criteria were: (1) contained data for patient-reported assessment of clinician empathy using the CARE Measure; and (2) provided CARE Measure data stratified by SES and/or race/ethnicity (including attempts to contact corresponding authors to obtain stratified data, when necessary). We considered studies eligible for inclusion regardless of language if the CARE Measure was previously validated in that language. We included both observational and interventional studies. We also included abstracts if they were published in a journal. We excluded studies for which stratified data could not be obtained. We also excluded studies that did not use the original CARE Measure (e.g. used an adaptation instead), and studies in which the CARE Measure was not completed by patients (e.g. completed by surrogates). We excluded editorials, correspondence, and review papers, as well as studies that were secondary reports of previously published studies.

### Search and identification of studies

We searched the electronic databases generally considered to be the most important sources^32^: Ovid MEDLINE, PubMed, CINAHL, EMBASE, CENTRAL and PsycINFO. We also performed a supplementary search of Google Scholar. Our previously published search strategy was as follows^31^:

Ovid MEDLINE (and adapted for searching the other databases)

1. “Consultation and Relational Empathy”.mp.
2. (CARE adj3 (measure* or question* or index*)).ti,ab. and empath*.mp.
3. (CARE adj3 (measure* or question* or index*)).ti,ab. and mercer.af.
4. 1 or 2 or 3

We adopted this search strategy and search terms from a previously conducted, comprehensive and rigorous systematic review of the CARE Measure.^28^ We consulted with a health librarian with expertise in systematic reviews who confirmed that the search strategy is methodologically sound. We searched from December 1, 2004 (date of the original publication of the CARE Measure) to present. We performed the search on May 21, 2020.

### Study selection and data abstraction

Two independent reviewers performed a relevance screen of the titles and abstracts of identified studies for potential eligibility. After the relevance screen, we compared the exclusion logs for the two reviewers and we calculated the Kappa statistic for assessment of interobserver agreement. In cases of disagreement, we reviewed the full manuscript for inclusion. All studies identified as potentially relevant in the relevance screen underwent full manuscript review. For each study that underwent full manuscript review, if the manuscript did not report stratified data (i.e. by SES and/or race/ethnicity) we sent an email query to the corresponding author to request stratified data. If there was no response to the initial request, we sent up to three follow-up author query emails approximately one week apart to request the data.

Using a standardized data collection form, two reviewers independently abstracted data for the following: (a) clinical context; (b) total number of patients; (c) definition of low SES (if applicable); (d) number of patients stratified by SES; (e) CARE Measure data stratified by SES (i.e. mean and standard deviation [SD]); (f) number of patients stratified by race/ethnicity; (g) CARE Measure data stratified by race/ethnicity (mean and SD). Any disagreements in the above processes were resolved by consensus with a third reviewer.

We abstracted and tabulated race/ethnicity data according to the race/ethnicity categories used in each of the included studies. To allow for pooling and comparing of data by race/ethnicity in a meta-analytic fashion we stratified abstracted data using the race/ethnicity categories for human subjects research recommended by the United States National Institutes of Health (NIH).^36^ For SES stratification, we adopted the definition of low SES used in each of the included manuscripts.

### Assessing study quality (risk of bias)

We used the Newcastle-Ottawa Scale to assess risk of bias as recommended in the Cochrane Handbook for cohort studies.^37,38^ If interventional studies were included, we also used the Newcastle-Ottawa Scale for risk of bias assessment because we are analyzing exposure (e.g. SES) and outcome (CARE Measure), and allocation/randomization are not relevant for what we are studying. We deemed studies to be low risk of bias if they had seven or more stars out of a possible nine stars on the Newcastle-Ottawa Scale.

### Analysis

As recommended in the Cochrane Handbook,^32^ we began with a qualitative analysis. We collated studies and summarized individual study results in table format. Where possible and appropriate, we pooled data and performed a quantitative analysis with a meta-analytic approach. As described in the protocol,^31^ because heterogeneous populations are needed in order to assess differences between race/ethnicity or SES groups, we only performed quantitative analysis for studies that had sufficient diversity in race/ethnicity and SES in the population (defined as no single race/ethnicity or SES group comprising >90% of the study population). We used separate random effects models to calculate pooled effect sizes and report mean differences with corresponding 95% confidence intervals (CIs) for low SES versus not low SES patients, as well as all non-white versus white patients. We also used separate random effects models to make pairwise comparisons (versus white patients) for Black/African American, Hispanic/Latino, Asian, and Native American patients.

We used the I^2^ statistic to assess heterogeneity in study results for each random effects model, with the following thresholds for interpretation: low heterogeneity: 25-49%; moderate heterogeneity: 50-74%; high heterogeneity: 75% or higher.^39^ We assessed for publication bias using funnel plots of the effect sizes against the precision of the studies.

Per our published protocol, we planned a sensitivity analysis restricted to studies with a low risk of bias as defined above. We also planned to analyze for possible interaction between SES and race/ethnicity, where possible, by comparing CARE Measure scores between SES categories stratified by race. We also performed post-hoc (i.e. not in our original protocol) analyses with meta-regression by year of publication, to test if there have been changes in empathy differences over time.

We used Stata 16 (StataCorp, College Station, TX) for all analyses.

## Results

Our database searches yielded 1085 records. After removal of duplicates, there were 748 independent studies that underwent relevance screen. Our Kappa calculation for the relevance screen was 0.83, indicating good inter-observer agreement. Following the relevance screen, 137 studies underwent full manuscript review. **Figure 1** displays the search, inclusion and exclusion of studies flow diagram.

**Figure 1:**
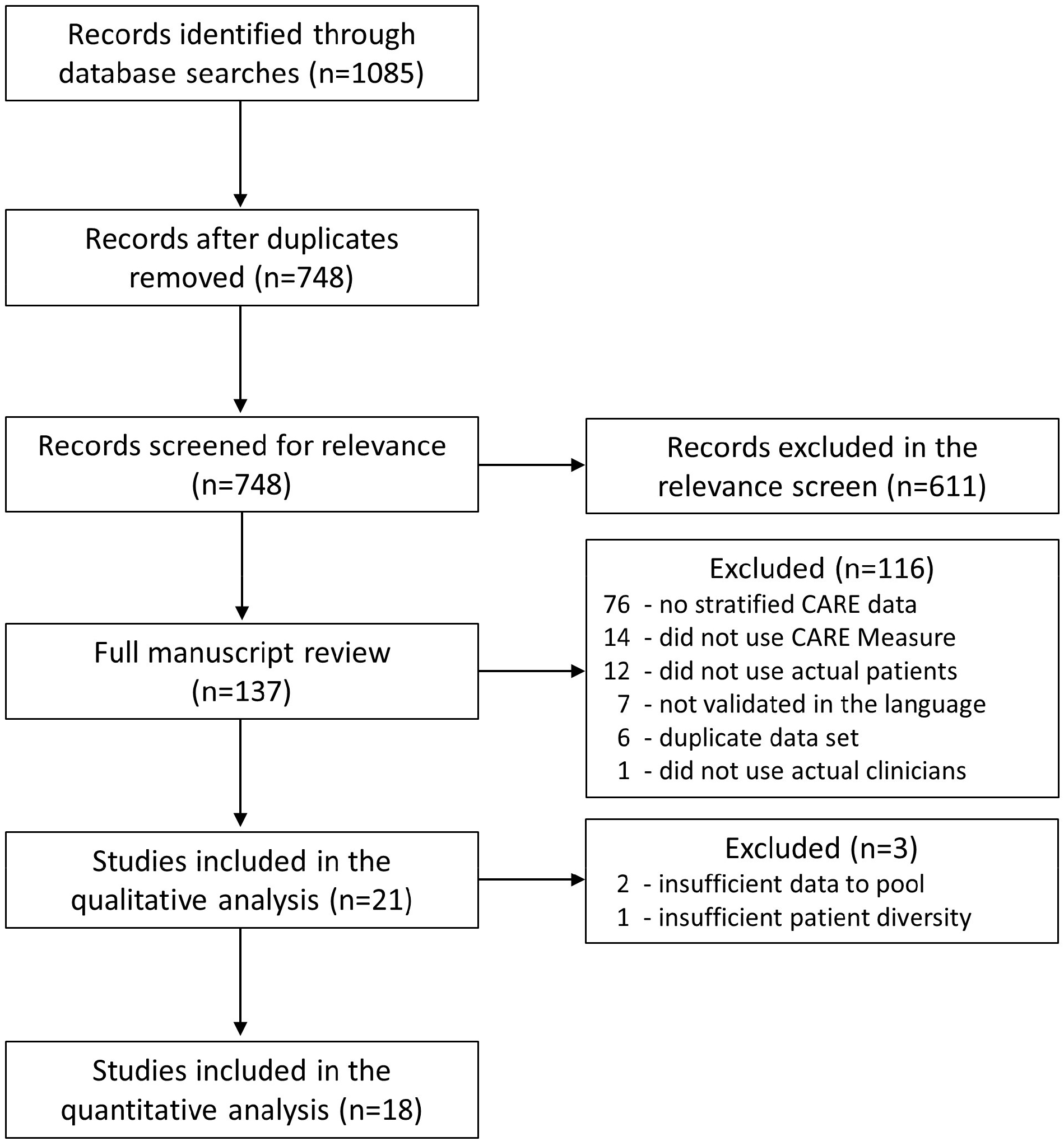
Search, inclusion and exclusion of studies flow diagram.

Twenty-one studies were included in the qualitative analyses. Three studies reported CARE Measure scores stratified by SES or race/ethnicity in the published manuscript, and the remaining 18 studies required queries to corresponding author to obtain stratified data. **Table 1** displays the nine studies with CARE Measure data stratified by SES.^40–48^ Most of the studies originated from the United Kingdom and the most common practice setting was primary care. **Table 2** displays the 14 studies with CARE Measure data stratified by race/ethnicity.^40,44,49-60^ Most of the studies were from the United States and the practice settings were diverse, including multidisciplinary practice, orthopedic surgery, emergency medicine, and primary care, among others. Only two studies had CARE Measure data stratified by both SES and race/ethnicity.^40,44^

**Table 1:** Studies containing CARE Measure data stratified by socioeconomic status.

| Author | PMID | Country | Context | Low SES Definition | Results, mean CARE score (SD) |
| --- | --- | --- | --- | --- | --- |
| Barrett | 21747102 | USA | Primary care | Annual income <\$25,000 (USD) | Low SES: n=163, 41.9 (5.6)<br>Not Low SES: n=517, 41.5 (6.2) |
| Bikker | 29204104 | UK | Sexual health practice | Unemployed and seeking work | Low SES: n=63, 47.8 (5.6)<br>Not Low SES: n=404, 48.1 (3.8) |
| Bikker | 26493072 | UK | Primary care | Unemployed and seeking work | Low SES: n=39, 44.8 (6.5)<br>Not Low SES: n=165, 45.9 (6.1) |
| Bikker | 16131282 | UK | Homeopathy practice | Postal codes | Low SES: n=12, 48.2 (2.9)<br>Not Low SES: n=25, 45.7 (5.5) |
| Hannan | 31182161 | USA | Multidisciplinary practice | Annual income <= \$49,400 (USD) | Low SES: n=56, 36.6 (10.9)<br>Not Low SES: n=125, 41.1 (8.6) |
| Jani | 22867682 | UK | Primary care | Scottish Index of Multiple Deprivation (lowest versus highest quartiles) | Low SES: n=107, 43.0 (6.8)<br>Not Low SES: n=56, 45.2 (6.8) |
| Mercer | 15772120 | UK | Primary care | Postal codes | Low SES: n=1832, 40.8 (9.0)<br>Not Low SES: n=2865, 40.9 (8.6) |
| Mercer | 26951586 | UK | Primary care | Scottish Index of Multiple Deprivation (lowest versus highest quartiles) | Low SES: n=356, 43.4 (6.6)<br>Not Low SES: n=302, 45.0 (6.2) |
| Yu | 26658427 | China | Multidisciplinary practice | Monthly income <\$5,000 (HKD) | Low SES: n=215, 33.0 (8.9)<br>Not Low SES: n=452, 34.8 (8.9) |

CARE: consultation and relational empathy; SES: socioeconomic status; PMID: PubMed identification number; SD: standard deviation; USA: United States of America; UK: United Kingdom; USD: United States dollars; HKD: Hong Kong dollars

**Table 2:**
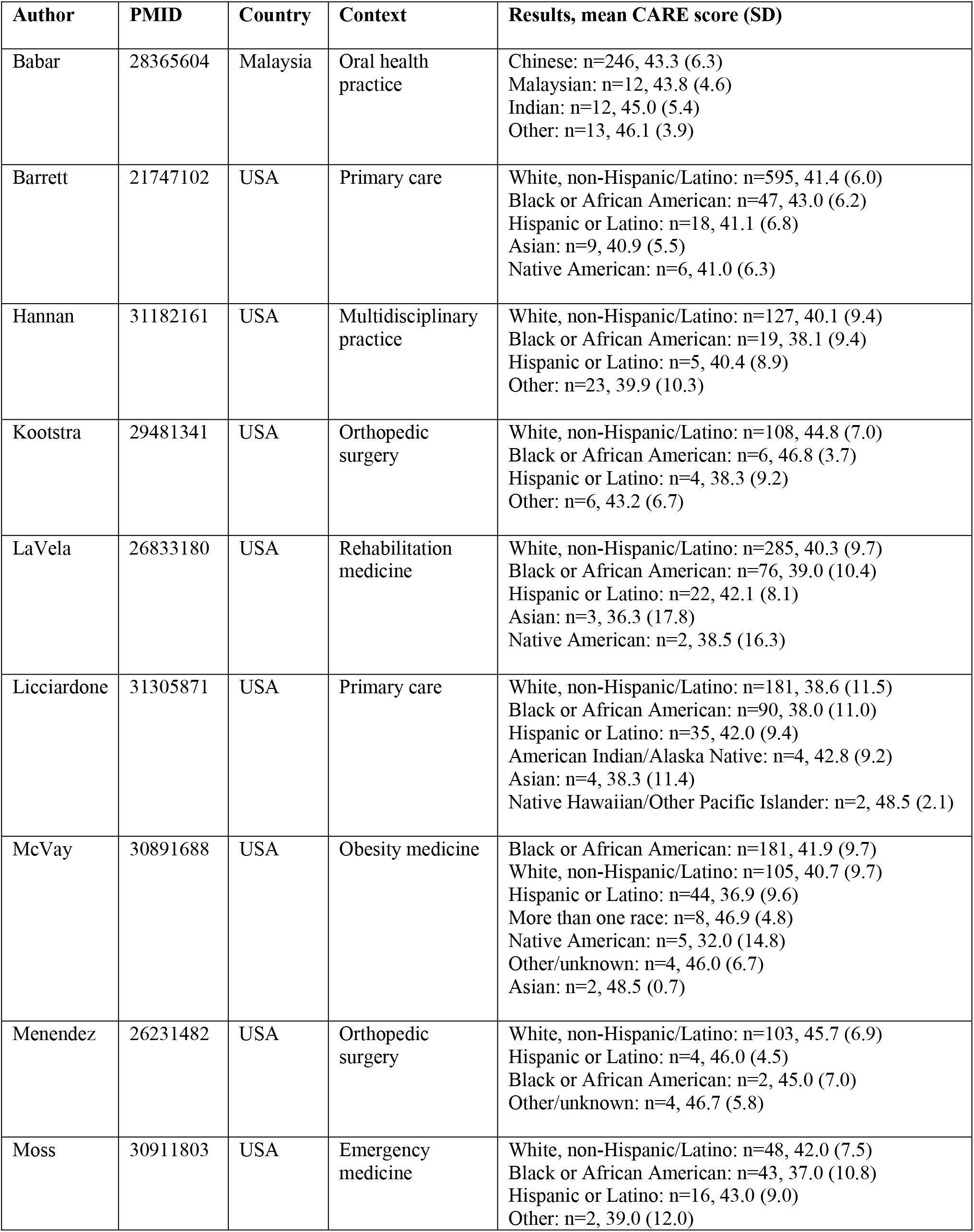
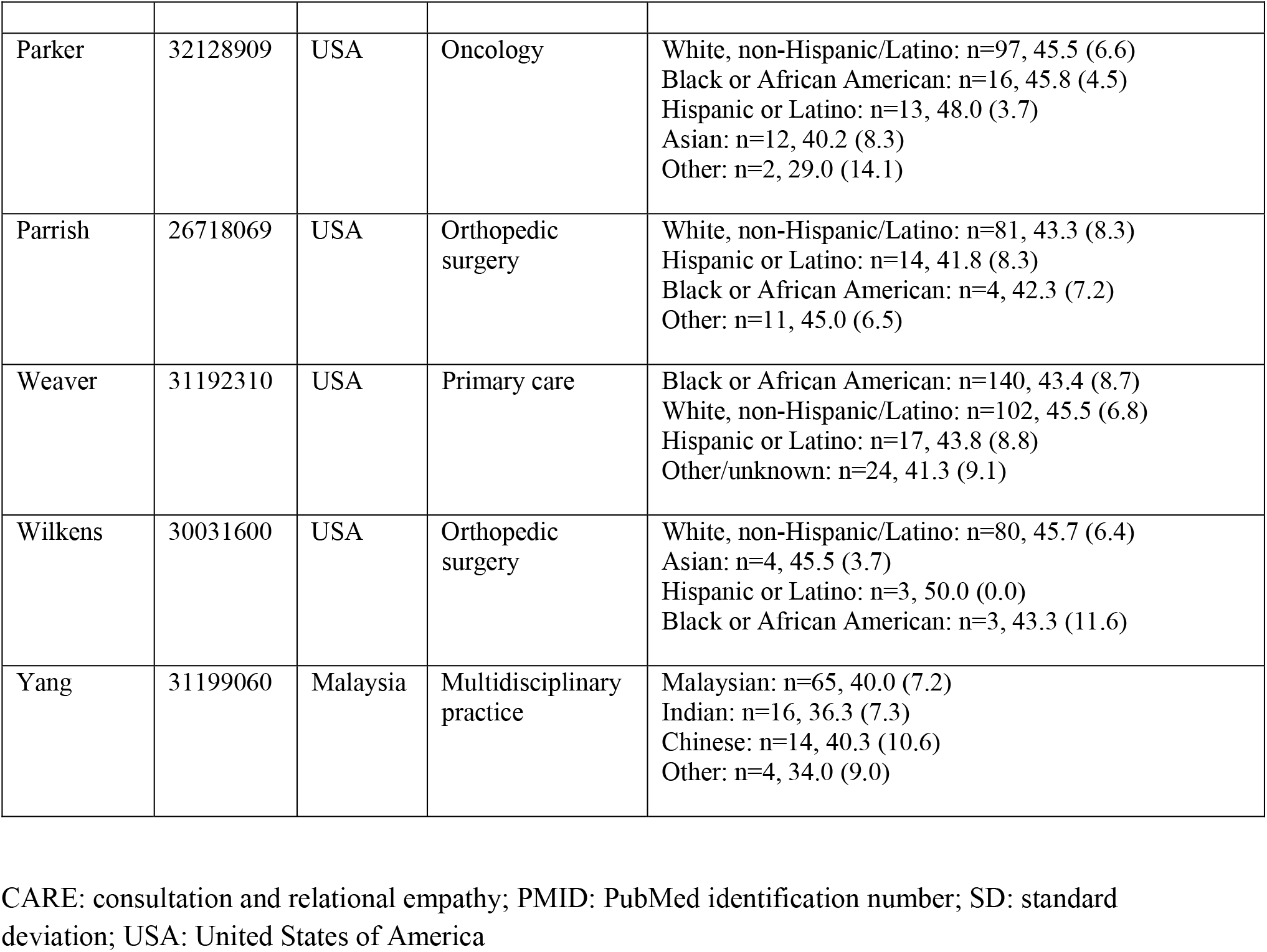
Studies containing CARE Measure data stratified by race/ethnicity.

| Author | PMID | Country | Context | Results, mean CARE score (SD) |
| --- | --- | --- | --- | --- |
| Babar | 28365604 | Malaysia | Oral health practice | Chinese: n=246, 43.3 (6.3)<br>Malaysian: n=12, 43.8 (4.6)<br>Indian: n=12, 45.0 (5.4)<br>Other: n=13, 46.1 (3.9) |
| Barrett | 21747102 | USA | Primary care | White, non-Hispanic/Latino: n=595, 41.4 (6.0)<br>Black or African American: n=47, 43.0 (6.2)<br>Hispanic or Latino: n=18, 41.1 (6.8)<br>Asian: n=9, 40.9 (5.5)<br>Native American: n=6, 41.0 (6.3) |
| Hannan | 31182161 | USA | Multidisciplinary practice | White, non-Hispanic/Latino: n=127, 40.1 (9.4)<br>Black or African American: n=19, 38.1 (9.4)<br>Hispanic or Latino: n=5, 40.4 (8.9)<br>Other: n=23, 39.9 (10.3) |
| Kootstra | 29481341 | USA | Orthopedic surgery | White, non-Hispanic/Latino: n=108, 44.8 (7.0)<br>Black or African American: n=6, 46.8 (3.7)<br>Hispanic or Latino: n=4, 38.3 (9.2)<br>Other: n=6, 43.2 (6.7) |
| LaVela | 26833180 | USA | Rehabilitation medicine | White, non-Hispanic/Latino: n=285, 40.3 (9.7)<br>Black or African American: n=76, 39.0 (10.4)<br>Hispanic or Latino: n=22, 42.1 (8.1)<br>Asian: n=3, 36.3 (17.8)<br>Native American: n=2, 38.5 (16.3) |
| Licciardone | 31305871 | USA | Primary care | White, non-Hispanic/Latino: n=181, 38.6 (11.5)<br>Black or African American: n=90, 38.0 (11.0)<br>Hispanic or Latino: n=35, 42.0 (9.4)<br>American Indian/Alaska Native: n=4, 42.8 (9.2)<br>Asian: n=4, 38.3 (11.4)<br>Native Hawaiian/Other Pacific Islander: n=2, 48.5 (2.1) |
| McVay | 30891688 | USA | Obesity medicine | Black or African American: n=181, 41.9 (9.7)<br>White, non-Hispanic/Latino: n=105, 40.7 (9.7)<br>Hispanic or Latino: n=44, 36.9 (9.6)<br>More than one race: n=8, 46.9 (4.8)<br>Native American: n=5, 32.0 (14.8)<br>Other/unknown: n=4, 46.0 (6.7)<br>Asian: n=2, 48.5 (0.7) |
| Menendez | 26231482 | USA | Orthopedic surgery | White, non-Hispanic/Latino: n=103, 45.7 (6.9)<br>Hispanic or Latino: n=4, 46.0 (4.5)<br>Black or African American: n=2, 45.0 (7.0)<br>Other/unknown: n=4, 46.7 (5.8) |
| Moss | 30911803 | USA | Emergency medicine | White, non-Hispanic/Latino: n=48, 42.0 (7.5)<br>Black or African American: n=43, 37.0 (10.8)<br>Hispanic or Latino: n=16, 43.0 (9.0)<br>Other: n=2, 39.0 (12.0) |
| Parker | 32128909 | USA | Oncology | White, non-Hispanic/Latino: n=97, 45.5 (6.6)<br>Black or African American: n=16, 45.8 (4.5)<br>Hispanic or Latino: n=13, 48.0 (3.7)<br>Asian: n=12, 40.2 (8.3)<br>Other: n=2, 29.0 (14.1) |
| Parrish | 26718069 | USA | Orthopedic surgery | White, non-Hispanic/Latino: n=81, 43.3 (8.3)<br>Hispanic or Latino: n=14, 41.8 (8.3)<br>Black or African American: n=4, 42.3 (7.2)<br>Other: n=11, 45.0 (6.5) |
| Weaver | 31192310 | USA | Primary care | Black or African American: n=140, 43.4 (8.7)<br>White, non-Hispanic/Latino: n=102, 45.5 (6.8)<br>Hispanic or Latino: n=17, 43.8 (8.8)<br>Other/unknown: n=24, 41.3 (9.1) |
| Wilkens | 30031600 | USA | Orthopedic surgery | White, non-Hispanic/Latino: n=80, 45.7 (6.4)<br>Asian: n=4, 45.5 (3.7)<br>Hispanic or Latino: n=3, 50.0 (0.0)<br>Black or African American: n=3, 43.3 (11.6) |
| Yang | 31199060 | Malaysia | Multidisciplinary practice | Malaysian: n=65, 40.0 (7.2)<br>Indian: n=16, 36.3 (7.3)<br>Chinese: n=14, 40.3 (10.6)<br>Other: n=4, 34.0 (9.0) |

CARE: consultation and relational empathy; PMID: PubMed identification number; SD: standard deviation; USA: United States of America

Eighteen studies were included in the quantitative meta-analyses. Of the three studies excluded from the quantitative meta-analysis, two were excluded because there was not enough data to pool (i.e. only two studies in a Malaysian population) and one was excluded because there was insufficient patient diversity in the sample as defined in the methods. The 18 studies in the meta-analysis included 9,708 patients in total, and 3,663 (38%) of the patients were either low SES or non-white.

**Figure 2** displays the results of the random effects model for SES. Overall, compared to patients with not low SES, low SES was associated with lower ratings of clinician empathy (mean CARE difference= -0.87 [95% CI -1.72 to -0.02]). While heterogeneity was moderate (I^2^=72%), none of the individual studies found significantly higher empathy for low SES patients.

**Figure 2:**
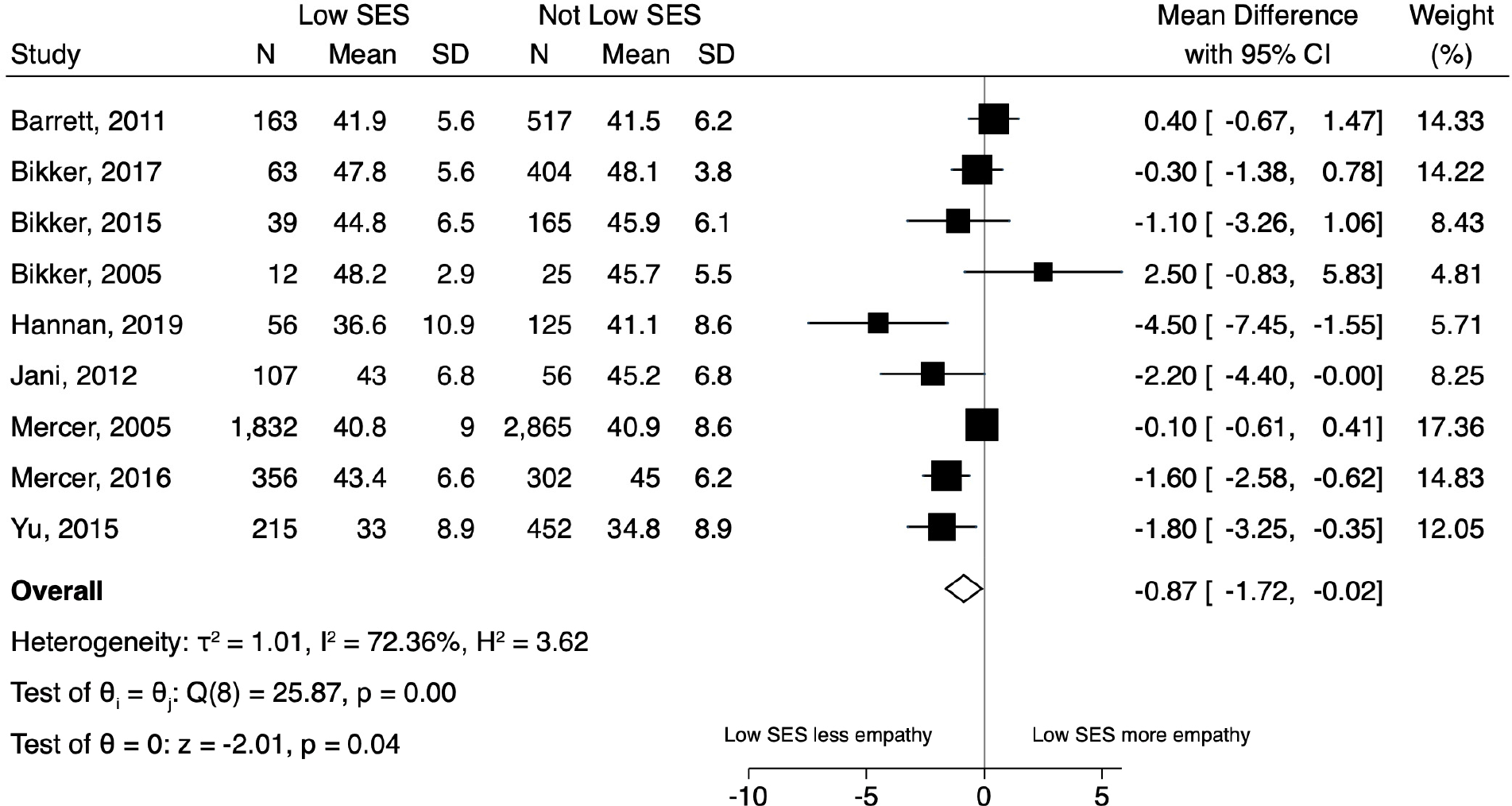
Forest plot of the results of the random effects model for patient-reported experience of clinician empathy comparing patients with low socioeconomic status (SES) to patients with not low SES.

**Figure 3** displays the results of the random effects model for all non-white patients compared to white patients. While we found that overall non-white patients reported lower clinician empathy compared to white patients, this difference was not statistically significant (mean CARE difference= -0.57 [95% CI -1.45 to 0.31]). The results of the separate (pairwise) random effects models for Black/African American, Hispanic/Latino, Asian, and Native American patients (compared to white patients) appear in the Electronic Supplementary Material (**eFigures 1-4**). In summary, compared to white patients, empathy scores were numerically lower for patients of multiple race/ethnicity groups (Black/African American, Asian, Native American, as well as all non-whites combined) but none of these differences reached statistical significance. Of note, in the five separate random effects models pertaining to race/ethnicity, none of the individual studies found significantly higher empathy for any group of non-whites.

**Figure 3:**
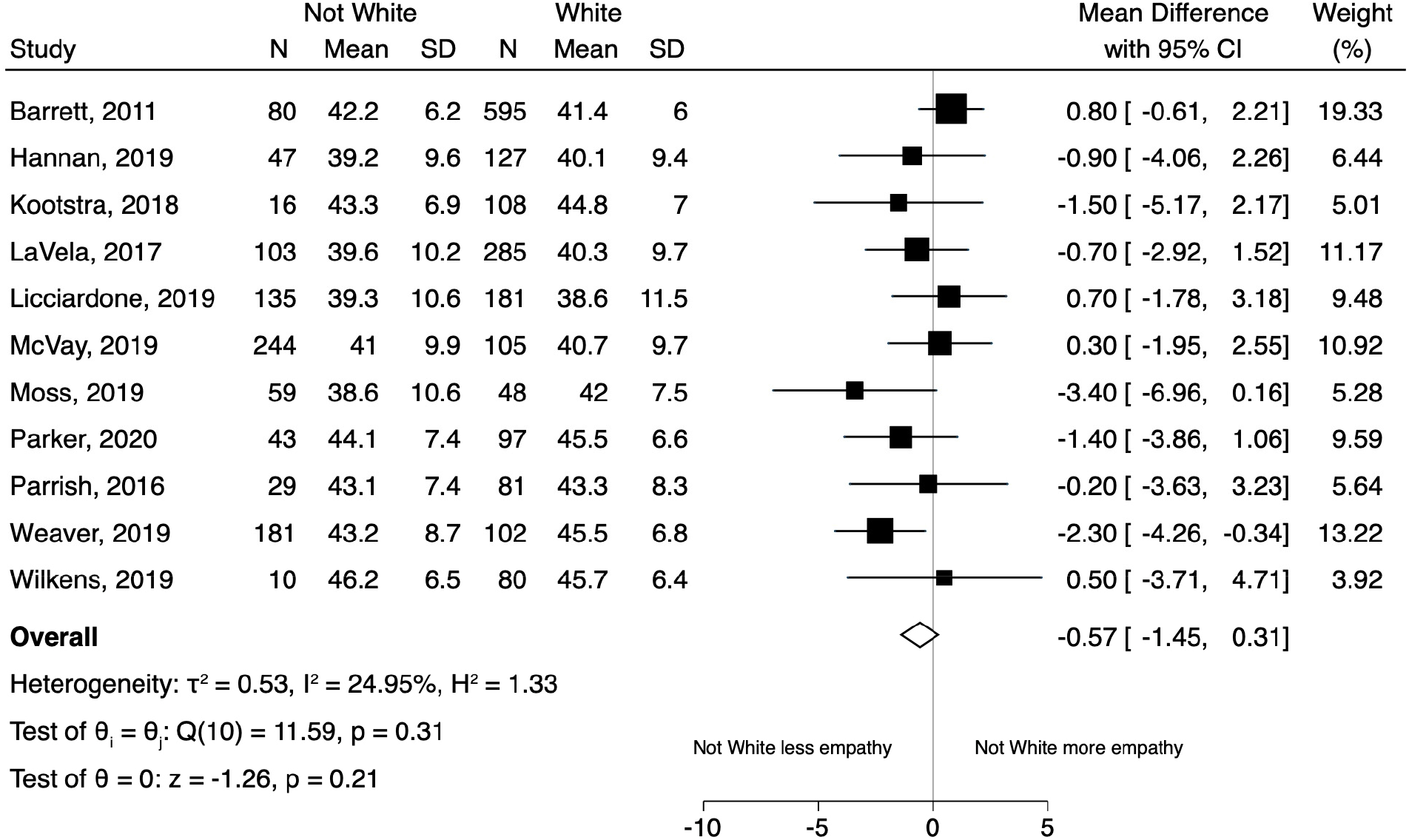
Forest plot of the results of the random effects model for patient-reported experience of clinician empathy comparing all non-white patients to white patients.

In the post-hoc meta-regression by year of publication, we found that empathy differences over time were increasing for both SES and race/ethnicity (for SES: -0.18 [95% CI -0.35 to -0.02] per year comparing low SES to not low SES; for race/ethnicity: -0.22 [95% CI -0.43 to -0.01] per year comparing non-white to white patients). These data suggest that empathy differences may be widening over time.

None of the included studies met our definition of low risk of bias; therefore, we were unable to perform the sensitivity analysis restricted to low risk of bias studies. Because only two of the studies had stratified data by both SES and race/ethnicity, we also were unable to analyze for possible interaction between SES and race/ethnicity. Visual inspection of the funnel plots for the SES and race/ethnicity analyses did not suggest publication bias (Electronic Supplementary Material, **eFigures 5-6**).

## Discussion

The purpose of this systematic review and meta-analysis was to generate preliminary data for testing the hypothesis that health care disparities exist in patient experience of clinician empathy (i.e. an empathy “gap”). After quantitatively analyzing 18 independent studies of the CARE Measure including more than nine thousand patients (and nearly 40% being low SES or non-white), we found that low SES patients had significantly lower patient-reported assessments of clinician empathy compared to patients with SES that was not low. Although we did not find statistically significant differences in empathy by race/ethnicity, we point to a trend that merits further research. The CARE Measure scores were consistently numerically lower for multiple race/ethnicity patient groups (compared to white patients), including Black/African American, Asian, Native American, and all non-whites combined. In addition, none of the included studies reported significantly higher empathy for any group of non-white patients compared to whites.

We believe there is some uniqueness in this report for two reasons. First, rather than testing for SES and race/ethnicity differences in patient satisfaction in the broad sense, we focused specifically on the element of empathy from clinicians. Second, although all health care disparities are likely rooted in a systemic lack of empathy for disadvantaged people (e.g. institutionalized racism), few data exist on differences in individual clinician empathy during face-to-face health care encounters (e.g. interpersonal racism).

We consider this work to be preliminary in nature given that there are important limitations to consider. First, the clinical significance of small differences in the CARE Measure are unclear, despite statistical significance (i.e. for SES). However, our post-hoc analyses identified that such an empathy gap may be widening over time. While exploratory in nature, these post-hoc analyses provide additional scientific rationale for future studies investigating the existence of an empathy gap, as well as studies aimed at increasing clinician empathy for disadvantaged populations. Further, given recent evidence that people from low SES communities are less likely to respond to patient experience surveys resulting in non-response bias (i.e. responders are not representative of the target patient population),^61^ future research needs to also focus on alternative methods to assess patient experience in disadvantaged populations so as to not underestimate possible differences in clinician empathy.

Another potential limitation is that our meta-analysis was limited to studies of the CARE Measure, and did not incorporate other previously published measures of clinician empathy. Our rationale was that we wanted to perform a quantitative meta-analysis, and this requires a single measure approach to limit heterogeneity and permit pooling of data. We selected the CARE Measure because to date it is the most commonly used assessment of clinician empathy from the patient perspective, and it has very well-validated methodology (i.e. proven reliability, internal validity and consistency).^28^ Nonetheless, it is possible that studies using a different measure would find different results.^62,63^

Importantly, as it pertains to the analyses by race/ethnicity, none of the studies contained information on race/ethnicity of the clinicians. Therefore, in our study it is not possible to account for race concordance/discordance (or in-group/out-group bias). We also acknowledge that the CARE Measure is a patient’s assessment of the empathy of clinicians (e.g. physicians) only. Others in the health care environment (e.g. clinic staff, registrars, etc.) may have interactions with patients that shape patients’ experience of empathy during health care encounters in a meaningful way, and this would not necessarily be captured by the CARE Measure. We also need to acknowledge that because individual patient-level data were not collected, we could not establish our own uniform definition of low SES, and instead we relied on the definition of low SES that the authors used in each individual study. Lastly, we are not aware of any studies that have tested if disadvantaged persons (by either SES and/or race/ethnicity) have different expectations for clinician empathy in health care encounters due to history of mistreatment (e.g. both institutionalized and interpersonal racism), and this could potentially affect the results.

Finding meaningful disparities in clinician empathy would have important implications for public health because clinician empathy is vital for high quality health care. We believe the results of this systematic review and meta-analysis are important preliminary data supporting that an empathy gap may exist for disadvantaged people in face-to-face health care encounters with clinicians. More research to further test this hypothesis and help promote health care equity is warranted.

## Data Availability

All of the studies included in this systematic review are already published in the public domain. After review and approval by our study data use committee, we will allow other researchers who submit to us a protocol to have unrestricted access to our database.

## Acknowledgements

We thank all of the corresponding authors of included studies who, in response to our author queries, provided stratified data (i.e. by SES and race/ethnicity) for analysis. We especially thank Prof. Stewart Mercer, as well as Shari Barlow, Dr. Bruce Barrett, Mary Checovich, Dr. Adriana Foster, Dr. Syed Shahzad Hasan, Dr. John Licciardone, Yi En Low, Wan Juen Ng, Dr. Raymond Parrish II, and Dr. Wai Yew Yang.

## Authors’ contributions

All authors have made substantial contributions to this paper and have satisfied the International Committee of Medical Journal Editors (ICMJE) criteria for authorship. BWR and ST supervised all aspects of the study and take responsibility for the paper as a whole. NKP and AJM contributed to the scientific approach and study design. NKP, CJT and ST conducted the search for identification of studies. NKP, CJT and ST abstracted and managed the data. CJT and ST sent the queries (data requests) to corresponding authors and managed the data. BWR and ST assessed study quality. BWR provided statistical expertise. ST drafted the manuscript. BWR, NKP, CJT and AJM contributed substantially to revision of the final manuscript. All authors approved the manuscript in its final form.

